# The cognitive phenotype of juvenile absence epilepsy and its heritability: An investigation of patients and unaffected siblings

**DOI:** 10.1101/2022.04.12.22273461

**Authors:** Lorenzo Caciagli, Corey Ratcliffe, Fenglai Xiao, Louis A. van Graan, Karin Trimmel, Christian Vollmar, Maria Centeno, John S. Duncan, Pamela J. Thompson, Sallie Baxendale, Matthias J. Koepp, Britta Wandschneider

## Abstract

**Objective:** The cognitive profile of juvenile absence epilepsy (JAE) remains uncharacterized. This study aimed to: (i) elucidate the neuropsychological profile of JAE; (ii) identify familial cognitive traits, by investigating unaffected JAE siblings; (iii) determine whether cognitive traits across the idiopathic generalized epilepsy (IGE) spectrum are shared or syndrome-specific, by comparing JAE to JME; and (iv) identify associations between cognitive abilities and clinical characteristics.

**Methods:** We investigated 123 participants: 23 patients with JAE, 16 unaffected siblings of JAE patients, 45 healthy controls, and 39 patients with JME, who underwent a comprehensive neuropsychological test battery including measures within four cognitive domains: attention/psychomotor speed, language, learning, and executive function. We also correlated clinical measures with cognitive performance data to decode effects of age at onset and duration of epilepsy.

**Results:** Patients with JAE performed worse than controls across tests of psychomotor speed, language, learning and executive function. Patients and siblings were similarly impaired on language measures of verbal comprehension, phonemic fluency, and semantic fluency compared to controls. Receiver operating characteristic curves indicated successful discrimination of patients with JAE and siblings from controls via linguistic measures. Individuals with JME also presented with multidomain cognitive impairment and had worse response inhibition than those with JAE. Across all patients, those with older age at onset had better performance on psychomotor speed and executive function tests.

**Significance:** JAE is associated with wide-ranging cognitive difficulties that encompass domains reliant on frontal lobe processing, including language, attention, and executive function. JAE siblings demonstrate shared impairment with patients on linguistic measures, indicative of a familial trait. Executive function subdomains may be differentially affected across the IGE spectrum. Cognitive abilities are detrimentally modulated by an early age at seizure onset.

**KEY POINTS:**

- JAE presents with multidomain cognitive impairment involving language, attention/ psychomotor speed, executive function, and learning.
- Impaired language is common to people with JAE and their unaffected siblings, suggestive of a familial trait (endophenotype).
- Response inhibition is worse in JME than JAE, indicating distinct cognitive profiles across the IGE spectrum.
- Early age at epilepsy onset is associated with worse cognitive performance in JAE and JME.

## INTRODUCTION

Juvenile absence epilepsy (JAE) is an idiopathic generalized epilepsy syndrome (IGE),^1^ and typically presents with onset of absence seizures in late childhood or adolescence. Most patients also experience generalized tonic-clonic seizures.^2^ JAE is assumed to be polygenetic in origin, similar to the other three IGE syndromes [childhood absence epilepsy (CAE), Juvenile Myoclonic Epilepsy (JME), and Generalized Tonic-Clonic Seizures Alone].^3^ Seizure onset in JAE and JME coincides with a crucial phase of neurodevelopment.^4^ It is hypothesized that alterations in developmental trajectories in JAE and JME may also lead to impaired cognition.^5–7^

Cognitive comorbidities are increasingly recognized as part of the IGE phenotype,^8–10^ can predate seizure onset by several years,^11,12^ and persist after seizure control is achieved.^9,13^ Cognitive difficulties have also been reported in seizure-unaffected first-degree relatives of patients with IGE and JME, the most common IGE syndrome.^14–16^ These cognitive traits are interpreted as intermediate phenotypes or *endophenotypes*, i.e., disease signatures that are more prevalent in patients and first-degree relatives than the general population, are closely related to the underlying genotype,^17^ and allow differentiating the heritable underpinnings of cognitive traits from the effects of disease activity or antiseizure medication (ASM).

Cognitive studies in absence epilepsies have focused on CAE or combined CAE and JAE cohorts, given the similarities in disease pathological mechanisms^18^ and clinical presentation,^2^ and revealed impairment of general intellectual abilities, visual-spatial processing, attention, language, and executive function.^8,19–21^ However, investigations that detail the cognitive profile of JAE, and probe the syndrome-specificity of cognitive traits, are still lacking.

Here, we aimed to characterize the cognitive phenotype of a homogeneous, well-defined JAE cohort via a comprehensive neuropsychological test battery. We also investigated unaffected siblings of JAE patients, to determine the heritability of cognitive profiles and identify JAE endophenotypes. Then, we directly compared individuals with JAE and JME, to highlight syndrome-specific and shared traits, and provide further insights into the presumed overlap of cognitive comorbidities across the IGE spectrum.^8^ Finally, we assessed the relationship between cognitive performance and clinical characteristics, such as age at onset and disease duration.

## METHODS

### Participants

In this prospective cross-sectional study, we investigated 123 consecutively recruited participants: 23 JAE patients, 16 seizure-unaffected siblings of 11 index patients with JAE, 39 JME patients, and 45 healthy control participants with no family history of epilepsy and other neurological disorders. All patients were recruited from epilepsy outpatient clinics at the National Hospital for Neurology and Neurosurgery (London, UK) and the Chalfont Centre for Epilepsy (Buckinghamshire, UK), between 2007 and 2019. Controls were recruited from local communities in North-West London and Chalfont St Peter, UK.

JAE patients had a typical clinical presentation, with age at onset in late childhood or early puberty [median (interquartile range, IQR) =12 (6) years]. All had absence seizures, and 83% had generalized tonic-clonic seizures (GTCS). Three patients (13%) reported infrequent myoclonus associated with absence seizures, which is compatible with a JAE diagnosis.^2^ Family history of epilepsy was confirmed in 12 patients (52%). All JAE patients had a typical routine EEG with interictal 3-4 Hz generalized spike-wave discharges; seven (30%) had been seizure-free for at least a year prior to the investigation. No JAE sibling had ever experienced seizures, except for one individual who had one clearly provoked GTCS episode following a head trauma during a motor vehicle accident. All JME patients had myoclonic seizures and GTCS, 14 of 39 patients (36%) had absences. All individuals with JME had a typical EEG with interictal generalized polyspike-and-wave discharges; twenty of them (51%) had been seizure-free for at least a year. Clinical MRIs were normal in all participants. We previously reported on some cognitive results of people with JME and part of the controls.^5,22^

Patients with JAE, their siblings and controls had comparable age. Patients with JME were older than those with JAE and controls. Groups were comparable for sex. Patients with JAE and their siblings had lower levels of education than controls. Related statistics, including further demographics and clinical details, are provided in Table 1.

**TABLE 1.** Demographic data, clinical characteristics, and questionnaires.

|  | **CTR**<br>**(n = 45)** | **JAE**<br>**(n = 23)** | **SIB**<br>**(n = 16)** | **JME**<br>**(n = 39)** | **Test Statistic** | **P value** | **Post-hoc tests**<br>**(Bonferroni**<br>**corrected)** |
| --- | --- | --- | --- | --- | --- | --- | --- |
| **Age, y, mean (SD)** | 28.4<br>(6.6) | 24.4<br>(6.6) | 26.0<br>(8.0) | 34.3<br>(10.7) | $F = 8.4$ | **<.001** | JAE vs CTR: .380<br>SIB vs CTR: 1.000<br>JAE vs SIB: 1.000<br>**JME vs JAE: <.001**<br>**JME vs CTR: .009** |
| **Sex, F/M** | 29/16 | 16/7 | 5/11 | 22/17 | $FET = 6.6$ | .079 | N/A |
| **Education, ordinal,**<br>**median (IQR)** | 3.0<br>(1.5) | 2.0<br>(2.0) | 2.0<br>(1.8) | 3.0<br>(1.0) | $H = 20.7$ | **<.001** | **JAE vs CTR: .001**<br>**SIB vs CTR: .001**<br>JAE vs SIB: 1.00<br>JME vs JAE: .834<br>JME vs CTR: .154 |
| **Dysexecutive Traits**<br>**(DEX) median (IQR)** | 14.0<br>(9.5) | 27.5<br>(14.8) | 10.0<br>(14.5) | 27.5<br>(18.5) | $H = 17.3$ | **<.001** | **JAE vs CTR: .008**<br>SIB vs CTR: 1.000<br>**JAE vs SIB: .010**<br>JME vs JAE: 1.000<br>JME vs CTR: .093 |
| **Anxiety**<br>**(HADS-A)**<br>**median (IQR)** | 5.0<br>(5.0) | 5.0<br>(6.0) | 2.5<br>(3.8) | 6.0<br>(5.5) | $H = 17.0$ | **<.001** | JAE vs CTR: .282<br>SIB vs CTR: .255<br>**JAE vs SIB: .004**<br>JME vs JAE: 1.00<br>JME vs CTR: .230 |
| **Depression**<br>**(HADS-D)**<br>**median (IQR)** | 1.0<br>(2.0) | 2.0<br>(3.0) | 0.0<br>(2.8) | 2.0<br>(4.3) | $H = 18.0$ | **<.001** | **JAE vs CTR: .021**<br>SIB vs CTR: 1.000<br>**JAE vs SIB: .030**<br>JME vs JAE: 1.00<br>JME vs CTR: .009 |
| **ASM, median (IQR)** | N/A | 2.0<br>(1.0) | N/A | 2.0<br>(1.0) | $H = .008$<br>(JME vs JAE) | .927 | |
| **Sodium valproate**<br>**(yes/no)** | N/A | | N/A | | $FET = 1.9$<br>(JME vs JAE) | .195 | |
| **Topiramate or**<br>**zonisamide (yes/no)** | N/A | 2/21 | N/A | 3/36 | $FET = .02$<br>(JME vs JAE) | .619 | |
| **Disease Duration, y,**<br>**median (IQR)** | N/A | 11.3<br>(11.0) | N/A | 16.0<br>(18.0) | $H = 5.9$<br>(JME vs JAE) | **.015** | |
| **Age at Onset,**<br>**y, median (IQR)** | - | 12.0<br>(6.0) | - | 15.0<br>(4.0) | $H = 9.3$<br>(JME vs JAE) | **.002** | |

*Abbreviations.* ASM= anti-seizure medication(s); CTR = Controls; FET = Fisher's Exact Test statistic; IQR= inter-quartile range; JAE = Juvenile Absence Epilepsy; JME = Juvenile Myoclonic Epilepsy; SD= standard deviation; SIB = Unaffected siblings of JAE patients.

### Standard protocol approvals, registrations, and patient consents

Participant recruitment received ethical approval from the University College London and University College London Hospitals Joint Research Ethics Committee (06/N059 and 11/LO/0439). Written informed consent was obtained from all participants in accordance with the standards of the Declaration of Helsinki.

### Self-assessment questionnaires

We used the Hospital Anxiety and Depression Scale (HADS), a self-assessment questionnaire, to address current symptoms of anxiety (HADS-A) and low mood (HADS-D).^23^ Participants also completed the Dysexecutive Questionnaire (DEX), which measures everyday life problems resulting from dysexecutive traits.^24^

### Neuropsychological tests

All participants underwent a comprehensive neuropsychological test battery, as described elsewhere,^8^ whose completion required about 90 minutes, with standardized interspersed breaks. Test details are provided in Supplementary Table 1.

General Intellectual level was estimated with the National Adult Reading Test (NART).^25^ In this study, we examined four cognitive domains: attention/psychomotor speed, language, learning, and executive function. Attention and psychomotor speed were assessed with the Trail Making Test (TMT, Part A)^26^ and Stroop Color-Word Test (Stroop C and W scores).^27^ Language tests included: (i) the Vocabulary subtest of the Wechsler Adult Intelligence Scale (WAIS) III edition,^28^ which addresses verbal comprehension and semantic knowledge; (ii) the WAIS Similarities subtest, which assesses verbal reasoning; (iii) the Controlled Oral Word Association Test (COWAT) for phonemic fluency;^8,29^ (iv) a category fluency test, employing “Animals”, “Fruits”, and “Vegetables” as categories;^8,29^ and (v) the McKenna Graded Naming Test, which assesses visual confrontation naming.^30^ Verbal learning, visual learning, verbal recall, and visual recall were measured with the List Learning and Design Learning Subtests from the Adult Memory and Information Processing Battery.^31^ Executive function tests included: (i) the Digit Span and (ii) Mental Arithmetic WAIS subtests, which both assess working memory;^28^ (iii) the Stroop Test (Colored Words– Interference measure),^27^ which assesses response inhibition; and (iv) the TMT Part B-A (Task Switching), which assesses mental flexibility.^26^ Letter fluency and similarities can also be regarded as executive function tests.^32^ Across all our participants, however, both letter fluency and similarities had higher correlations with language than with executive function measures (Supplementary Methods), and were thus considered as language measures.^33^

### Statistical analyses

Data were analyzed using IBM SPSS v.28 and R 4.0.3. For analysis of demographic and clinical data, we used ANOVA, Kruskal-Wallis, and Fisher’s exact tests for continuous parametric, non-parametric and categorical data, respectively; *post-hoc* tests were Bonferroni-corrected for multiple comparisons. Kruskal-Wallis and Fisher’s exact test were used to compare clinical parameters between individuals with JAE and JME. Some data for education, questionnaires, and some cognitive tests were missing because of slight changes in the study protocol. Thus, we used Little’s missing completely at random (MCAR)^34^ on all cases, all neuropsychological test measures, and all education, anxiety, depression, and dysexecutive trait questionnaires, which showed no association between data missingness and any values (χ^2^ =506.8, *df*=480, *p*=.19).

We used multivariate analysis of covariance (MANCOVA) to compare neuropsychological test data among groups. In all models, we employed raw test scores, included age and (binary) sex as covariates, and used Wilk’s lambda (λ) as multivariate test statistic. To mitigate the risk for redundancy among the dependent variables of the MANCOVAs, we included only one measure from the Stroop test, the Interference score. To address our study aims, we envisioned the following models:

i. comparison of individuals with JAE, their unaffected siblings, and controls, to ascertain the cognitive profile of JAE and identify JAE endophenotypes;
ii. sensitivity analyses, consisting of model (i) rerun with additional covariates, represented by features that differed across groups: education (first sensitivity analysis), and depression and anxiety (second sensitivity analysis);
iii. comparison of the JAE subgroup with uncontrolled seizures (n=16) and healthy controls, to address the potential influence of disease severity;
iv. comparison of JAE and JME, to address the syndrome specificity of cognitive profiles;
v. for completeness, comparison of individuals with JME and healthy controls.

For all neuropsychological tests, we then performed univariate analyses via ANCOVAs, as previously,^5,16^ and compared: (i) JAE patients, JAE siblings, and controls (both in main models and sensitivity analyses); (ii) JAE patients with ongoing seizures versus controls; (iii) JAE patients versus JME patients. For completeness, we also compared (iv) JME patients versus controls. For these ‘test-wise’ ANCOVAs, we used age and sex as covariates, and adjusted *p*-values of each test for multiple comparisons via the false discovery rate (FDR) procedure; all *post-hoc* pairwise tests were Bonferroni-corrected. Cohen’s *d* was used as a measure of effect size. In all figures, test scores are plotted as raw, while the shown *p*-values refer to age- and sex-adjusted statistics. Missing data were addressed via pairwise deletion.

### Cognitive endophenotypes of JAE: individualized participant discrimination

We used a subset of tests yielding significant differences among JAE patients, siblings, and controls (verbal comprehension, phonemic fluency, and semantic fluency) for receiver operating characteristic (ROC) curves analyses, which assess accuracy of individual-level discrimination, and further validate neuropsychological test measures as JAE endophenotypes. We first adjusted cognitive scores for age and sex via linear regression, and then used unstandardized residuals for ROC curve analyses. For all measures, we assessed (i) discrimination of individuals with JAE from controls, and (ii) discrimination of individuals in a combined group of patients and their siblings from controls, using the area under the curve (AUC) metric.^5^

### Correlation analyses

We correlated cognitive test scores across all patients with age at onset, to probe the influence of timing of disease onset on cognition. We also conducted correlation analyses of cognitive measures with disease duration as a marker of disease chronicity. Firstly, we ran three Principal Components Analyses (PCA) to reduce data dimensionality,^5^ and obtained composite cognitive constructs for: (i) *attention/psychomotor speed* (Trail Making A, Stroop Words, Stroop Colors); (ii) *language* (graded naming test, phonemic fluency, semantic fluency, Vocabulary, and Similarities); and (iii) *executive function* (Digit span, Arithmetic, Trail Making Test B-A, and Stroop response inhibition). For each PCA, we verified that the first principal component (PC) had an eigenvalue > 1 and explained a sizeable amount of variance (>40%). We retained the first PC for each cognitive construct for correlation analyses. As the correlation between age at onset and disease duration approached statistical significance (*ρ*=-.24, *p*=.068), we opted for partial correlations of PCs with age at onset, covaried for duration, and vice versa. Chronological age and age at onset were not significantly correlated (*ρ*=-.10, *p*=.46).

### Data availability statement

Anonymized statistical data are available from the corresponding authors upon reasonable request from any qualified investigator.

## RESULTS

### Demographic, clinical data and self-assessment questionnaires

Complete statistical details are provided in Table 1. Patients with JAE reported more symptoms of depression than their siblings and controls (*p*_Bonferroni_ =.030/.021, respectively), and more symptoms of anxiety than their siblings (*p*_Bonferroni_ =.004). Median scores for anxiety and depression symptoms were largely smaller than cut-off scores used to define mild symptoms in all groups.^23^ Self-reported dysexecutive traits were more pronounced in individuals with JAE than in siblings and controls (*p*_Bonferroni_ =.010/.008, respectively). Patient groups were comparable for ASM number, proportion of patients treated with sodium valproate, and proportion of patients treated with topiramate or zonisamide. Patients with JAE had younger age at onset and shorter disease duration than those with JME (*p=*.002/.015, respectively).

### Cognitive performance in JAE, JAE siblings, and controls

Statistical details are provided in Table 2. MANCOVA yielded a significant effect of group for cognitive performance (Wilk’s λ28, *F*_(30,70)_=2.11, *p*=.005). Follow-up ANCOVAs showed significant group differences across multiple cognitive domains, including language (vocabulary, similarities, phonemic and semantic fluency; Figure 1A, 1B), attention and psychomotor speed (Stroop–Words, TMT-A), executive function (arithmetic, TMT B-A; Figure 2A-C) and learning (list learning, design learning and recall; Figure 3A, 3B; for all the above, *p*_FDR_<.05, Table 2). *Post-hoc* tests showed worse performance in JAE than controls on all the above listed tests except for verbal learning (all *p*_Bonferroni_ ≤ .025; Cohen’s *d* range: |0.75| to |1.32|). Individuals with JAE and their siblings had overlapping language impairment, as indicated by measures of vocabulary, phonemic and semantic fluency (all *p*_Bonferroni_ ≤.01 versus controls; *d*, vocabulary/phonemic fluency/semantic fluency= -1.10/-1.10/-1.00 for JAE, and - 1.06/-1.13/-1.22 for siblings, respectively). For the remaining tests, JAE siblings had an intermediate position between patients and controls, and no statistically significant differences against either group.

**TABLE 2.**
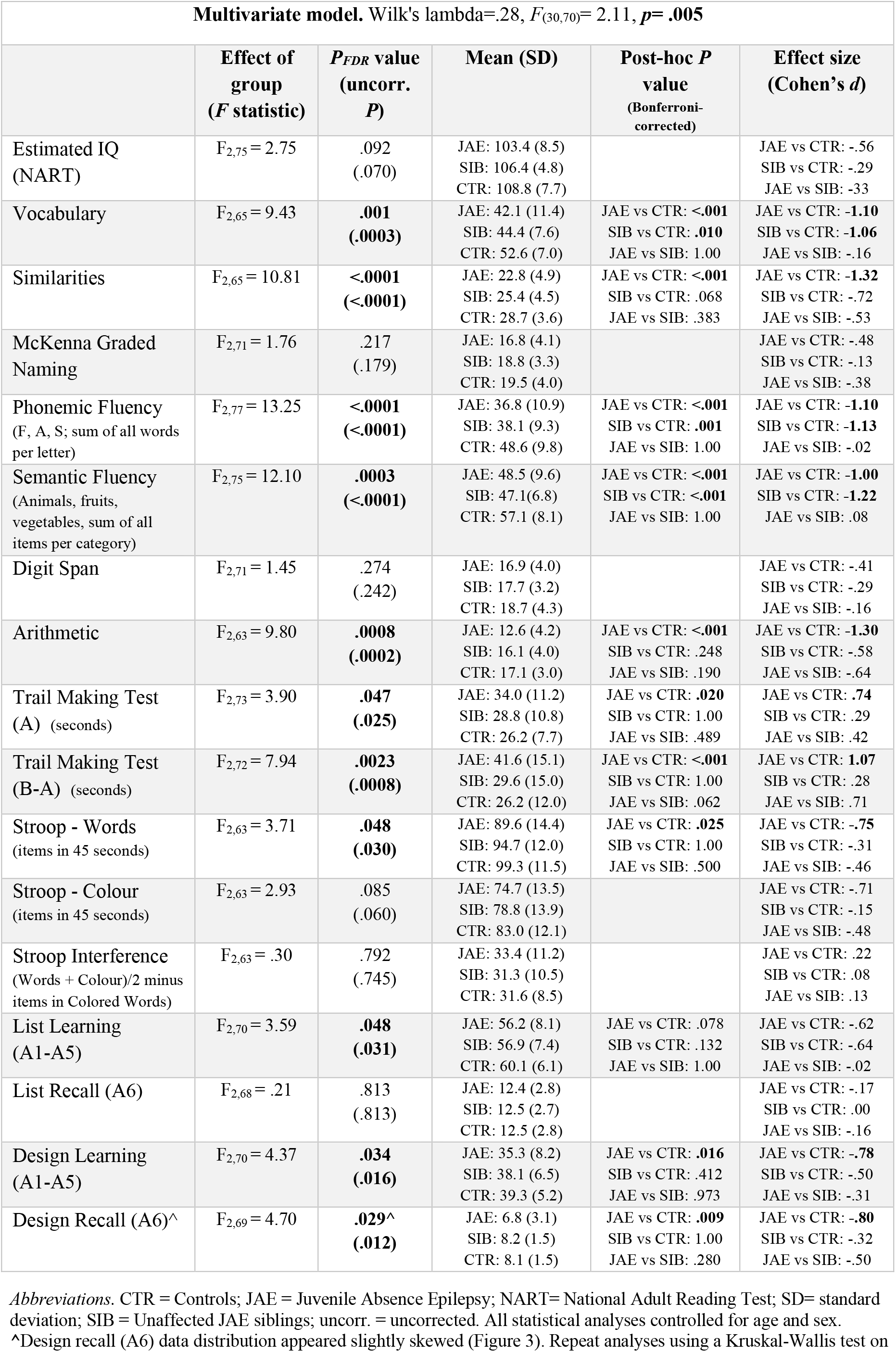
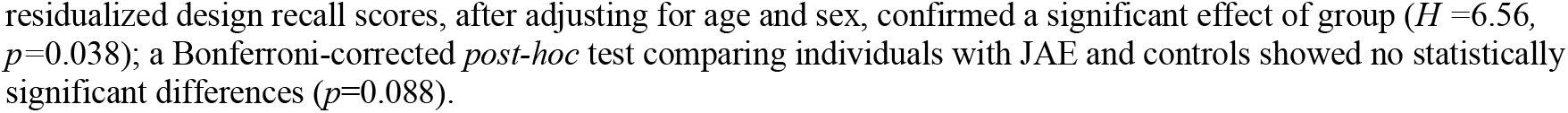
Comparison of JAE, JAE siblings, and controls.

| Multivariate model. Wilk's lambda=.28, $F_{(30,70)} = 2.11$, $p = .005$ | | | | | |
| --- | --- | --- | --- | --- | --- |
|  | Effect of group<br>(*F* statistic) | *P*<sub>FDR</sub> value<br>(uncorr.<br>*P*) | Mean (SD) | Post-hoc *P*<br>value<br>(Bonferroni-corrected) | Effect size<br>(Cohen's *d*) |
| Estimated IQ<br>(NART) | $F_{2,75} = 2.75$ | .092<br>(.070) | JAE: 103.4 (8.5)<br>SIB: 106.4 (4.8)<br>CTR: 108.8 (7.7) | | JAE vs CTR: -.56<br>SIB vs CTR: -.29<br>JAE vs SIB: -.33 |
| Vocabulary | $F_{2,65} = 9.43$ | **.001**<br>**(.0003)** | JAE: 42.1 (11.4)<br>SIB: 44.4 (7.6)<br>CTR: 52.6 (7.0) | JAE vs CTR: <**.001**<br>SIB vs CTR: **.010**<br>JAE vs SIB: 1.00 | JAE vs CTR: **-1.10**<br>SIB vs CTR: **-1.06**<br>JAE vs SIB: -.16 |
| Similarities | $F_{2,65} = 10.81$ | **<.0001**<br>**(<.0001)** | JAE: 22.8 (4.9)<br>SIB: 25.4 (4.5)<br>CTR: 28.7 (3.6) | JAE vs CTR: <**.001**<br>SIB vs CTR: .068<br>JAE vs SIB: .383 | JAE vs CTR: **-1.32**<br>SIB vs CTR: -.72<br>JAE vs SIB: -.53 |
| McKenna Graded<br>Naming | $F_{2,71} = 1.76$ | .217<br>(.179) | JAE: 16.8 (4.1)<br>SIB: 18.8 (3.3)<br>CTR: 19.5 (4.0) | | JAE vs CTR: -.48<br>SIB vs CTR: -.13<br>JAE vs SIB: -.38 |
| Phonemic Fluency<br>(F, A, S; sum of all words<br>per letter) | $F_{2,77} = 13.25$ | **<.0001**<br>**(<.0001)** | JAE: 36.8 (10.9)<br>SIB: 38.1 (9.3)<br>CTR: 48.6 (9.8) | JAE vs CTR: <**.001**<br>SIB vs CTR: **.001**<br>JAE vs SIB: 1.00 | JAE vs CTR: **-1.10**<br>SIB vs CTR: **-1.13**<br>JAE vs SIB: -.02 |
| Semantic Fluency<br>(Animals, fruits,<br>vegetables, sum of all<br>items per category) | $F_{2,75} = 12.10$ | **.0003**<br>**(<.0001)** | JAE: 48.5 (9.6)<br>SIB: 47.1 (6.8)<br>CTR: 57.1 (8.1) | JAE vs CTR: <**.001**<br>SIB vs CTR: <**.001**<br>JAE vs SIB: 1.00 | JAE vs CTR: **-1.00**<br>SIB vs CTR: **-1.22**<br>JAE vs SIB: .08 |
| Digit Span | $F_{2,71} = 1.45$ | .274<br>(.242) | JAE: 16.9 (4.0)<br>SIB: 17.7 (3.2)<br>CTR: 18.7 (4.3) | | JAE vs CTR: -.41<br>SIB vs CTR: -.29<br>JAE vs SIB: -.16 |
| Arithmetic | $F_{2,63} = 9.80$ | **.0008**<br>**(.0002)** | JAE: 12.6 (4.2)<br>SIB: 16.1 (4.0)<br>CTR: 17.1 (3.0) | JAE vs CTR: <**.001**<br>SIB vs CTR: .248<br>JAE vs SIB: .190 | JAE vs CTR: **-1.30**<br>SIB vs CTR: -.58<br>JAE vs SIB: -.64 |
| Trail Making Test<br>(A) (seconds) | $F_{2,73} = 3.90$ | **.047**<br>**(.025)** | JAE: 34.0 (11.2)<br>SIB: 28.8 (10.8)<br>CTR: 26.2 (7.7) | JAE vs CTR: **.020**<br>SIB vs CTR: 1.00<br>JAE vs SIB: .489 | JAE vs CTR: **.74**<br>SIB vs CTR: .29<br>JAE vs SIB: .42 |
| Trail Making Test<br>(B-A) (seconds) | $F_{2,72} = 7.94$ | **.0023**<br>**(.0008)** | JAE: 41.6 (15.1)<br>SIB: 29.6 (15.0)<br>CTR: 26.2 (12.0) | JAE vs CTR: <**.001**<br>SIB vs CTR: 1.00<br>JAE vs SIB: .062 | JAE vs CTR: **1.07**<br>SIB vs CTR: .28<br>JAE vs SIB: .71 |
| Stroop - Words<br>(items in 45 seconds) | $F_{2,63} = 3.71$ | **.048**<br>**(.030)** | JAE: 89.6 (14.4)<br>SIB: 94.7 (12.0)<br>CTR: 99.3 (11.5) | JAE vs CTR: **.025**<br>SIB vs CTR: 1.00<br>JAE vs SIB: .500 | JAE vs CTR: **-.75**<br>SIB vs CTR: -.31<br>JAE vs SIB: -.46 |
| Stroop - Colour<br>(items in 45 seconds) | $F_{2,63} = 2.93$ | .085<br>(.060) | JAE: 74.7 (13.5)<br>SIB: 78.8 (13.9)<br>CTR: 83.0 (12.1) | | JAE vs CTR: -.71<br>SIB vs CTR: -.15<br>JAE vs SIB: -.48 |
| Stroop Interference<br>(Words + Colour)/2 minus<br>items in Colored Words) | $F_{2,63} = .30$ | .792<br>(.745) | JAE: 33.4 (11.2)<br>SIB: 31.3 (10.5)<br>CTR: 31.6 (8.5) | | JAE vs CTR: .22<br>SIB vs CTR: .08<br>JAE vs SIB: .13 |
| List Learning<br>(A1-A5) | $F_{2,70} = 3.59$ | **.048**<br>**(.031)** | JAE: 56.2 (8.1)<br>SIB: 56.9 (7.4)<br>CTR: 60.1 (6.1) | JAE vs CTR: .078<br>SIB vs CTR: .132<br>JAE vs SIB: 1.00 | JAE vs CTR: -.62<br>SIB vs CTR: -.64<br>JAE vs SIB: -.02 |
| List Recall (A6) | $F_{2,68} = .21$ | .813<br>(.813) | JAE: 12.4 (2.8)<br>SIB: 12.5 (2.7)<br>CTR: 12.5 (2.8) | | JAE vs CTR: -.17<br>SIB vs CTR: .00<br>JAE vs SIB: -.16 |
| Design Learning<br>(A1-A5) | $F_{2,70} = 4.37$ | **.034**<br>**(.016)** | JAE: 35.3 (8.2)<br>SIB: 38.1 (6.5)<br>CTR: 39.3 (5.2) | JAE vs CTR: **.016**<br>SIB vs CTR: .412<br>JAE vs SIB: .973 | JAE vs CTR: **-.78**<br>SIB vs CTR: -.50<br>JAE vs SIB: -.31 |
| Design Recall (A6)^ | $F_{2,69} = 4.70$ | **.029^**<br>**(.012)** | JAE: 6.8 (3.1)<br>SIB: 8.2 (1.5)<br>CTR: 8.1 (1.5) | JAE vs CTR: **.009**<br>SIB vs CTR: 1.00<br>JAE vs SIB: .280 | JAE vs CTR: **-.80**<br>SIB vs CTR: -.32<br>JAE vs SIB: -.50 |

*Abbreviations.* CTR = Controls; JAE = Juvenile Absence Epilepsy; NART= National Adult Reading Test; SD= standard deviation; SIB = Unaffected JAE siblings; uncorr. = uncorrected. All statistical analyses controlled for age and sex.
^Design recall (A6) data distribution appeared slightly skewed (Figure 3). Repeat analyses using a Kruskal-Wallis test on
residualized design recall scores, after adjusting for age and sex, confirmed a significant effect of group ( $H = 6.56$ , $p = 0.038$ ); a Bonferroni-corrected *post-hoc* test comparing individuals with JAE and controls showed no statistically significant differences ( $p = 0.088$ ).

**TABLE 3.** Comparison of JAE and JME.

| *Multivariate model:* Wilks's lambda = .44, $F_{(15,24)} = 2.08$, $p = .053$ | | | | |
| --- | --- | --- | --- | --- |
|  | Effect of group<br>(*F* statistic) | *P*<sub>FDR</sub> value<br>(uncorr. *P*) | Mean (SD) | Effect size<br>(Cohen's *d*) |
| Estimated IQ (NART) | $F_{1,54} = 2.14$ | .633<br>(.149) | JAE: 103.4 (8.5)<br>JME: 108.4 (10.9) | JAE vs JME: -.35 |
| Vocabulary | $F_{1,54} = .48$ | .823<br>(.494) | JAE: 42.1 (11.4)<br>JME: 48.1 (9.5) | JAE vs JME: -.17 |
| Similarities | $F_{1,54} = .38$ | .823<br>(.538) | JAE: 22.8 (4.9)<br>JME: 23.6 (4.4) | JAE vs JME: .15 |
| McKenna | $F_{1,56} = .24$ | .823<br>(.629) | JAE: 16.8 (4.1)<br>JME: 18.3 (3.9) | JAE vs JME: .12 |
| Phonemic Fluency<br>(F, A, S; sum of all words per<br>letter) | $F_{1,57} = .31$ | .823<br>(.579) | JAE: 36.8 (10.9)<br>JME: 41.1 (11.08) | JAE vs JME: -.13 |
| Semantic Fluency<br>(Animals, fruits, vegetables, sum<br>of all items per category) | $F_{1,57} = 1.62$ | .707<br>(.208) | JAE: 48.5 (9.6)<br>JME: 52.2 (12.4) | JAE vs JME: -.31 |
| Digit Span | $F_{1,54} = .32$ | .823<br>(.577) | JAE: 16.9 (4.0)<br>JME: 19.0 (4.4) | JAE vs JME: -.14 |
| Arithmetic | $F_{1,48} = .00$ | .986<br>(.949) | JAE: 12.6 (4.2)<br>JME: 14.3 (4.2) | JAE vs JME: -.02 |
| Trail Making Test (A)<br>(seconds) | $F_{1,57} = .34$ | .823<br>(.564) | JAE: 34.0 (11.2)<br>JME: 31.3 (10.3) | JAE vs JME: .14 |
| Trail Making Test (B-A)<br>(seconds) | $F_{1,57} = .42$ | .823<br>(.520) | JAE: 34.0 (11.2)<br>JME: 37.4 (16.9) | JAE vs JME: .16 |
| Stroop Words<br>(items in 45 seconds) | $F_{1,52} = 3.09$ | .482<br>(.085) | JAE: 89.6 (14.4)<br>JME: 99.7 (20.2) | JAE vs JME: -.43 |
| Stroop Colour<br>(items in 45 seconds) | $F_{1,52} = .273$ | .823<br>(.603) | JAE: 74.7 (13.5)<br>JME: 74.3 (14.9) | JAE vs JME: -.13 |
| Stroop Interference<br>(Words + Colour)/2 minus items in<br>Colored Words) | $F_{1,52} = 6.36$ | .256<br>**(.015)** | JAE: 33.4 (11.2)<br>JME: 43.0 (13.4) | JAE vs JME: **.62** |
| List Learning (A1-A5) | $F_{1,56} = .02$ | .986<br>(.887) | JAE: 56.2 (8.1)<br>JME: 54.2 (8.6) | JAE vs JME: .04 |
| List Recall (A6) | $F_{1,55} = .00$ | .986<br>(.978) | JAE: 12.4 (2.8)<br>JME: 11.6 (2.6) | JAE vs JME: -.01 |
| Design Learning (A1-A5) | $F_{1,54} = .00$ | .986<br>(.986) | JAE: 35.3 (8.2)<br>JME: 34.8 (8.9) | JAE vs JME: .01 |
| Design Recall (A6) | $F_{1,54} = 3.30$ | .482<br>(.075) | JAE: 6.8 (3.1)<br>JME: 7.4 (2.0) | JAE vs JME: -.46 |

*Abbreviations.* JAE = Juvenile Absence Epilepsy; JME = Juvenile Myoclonic Epilepsy; NART = National Adult Reading Test; SD = standard deviation; uncorr. = uncorrected. All statistical analyses controlled for age and sex.

**Figure 1.**
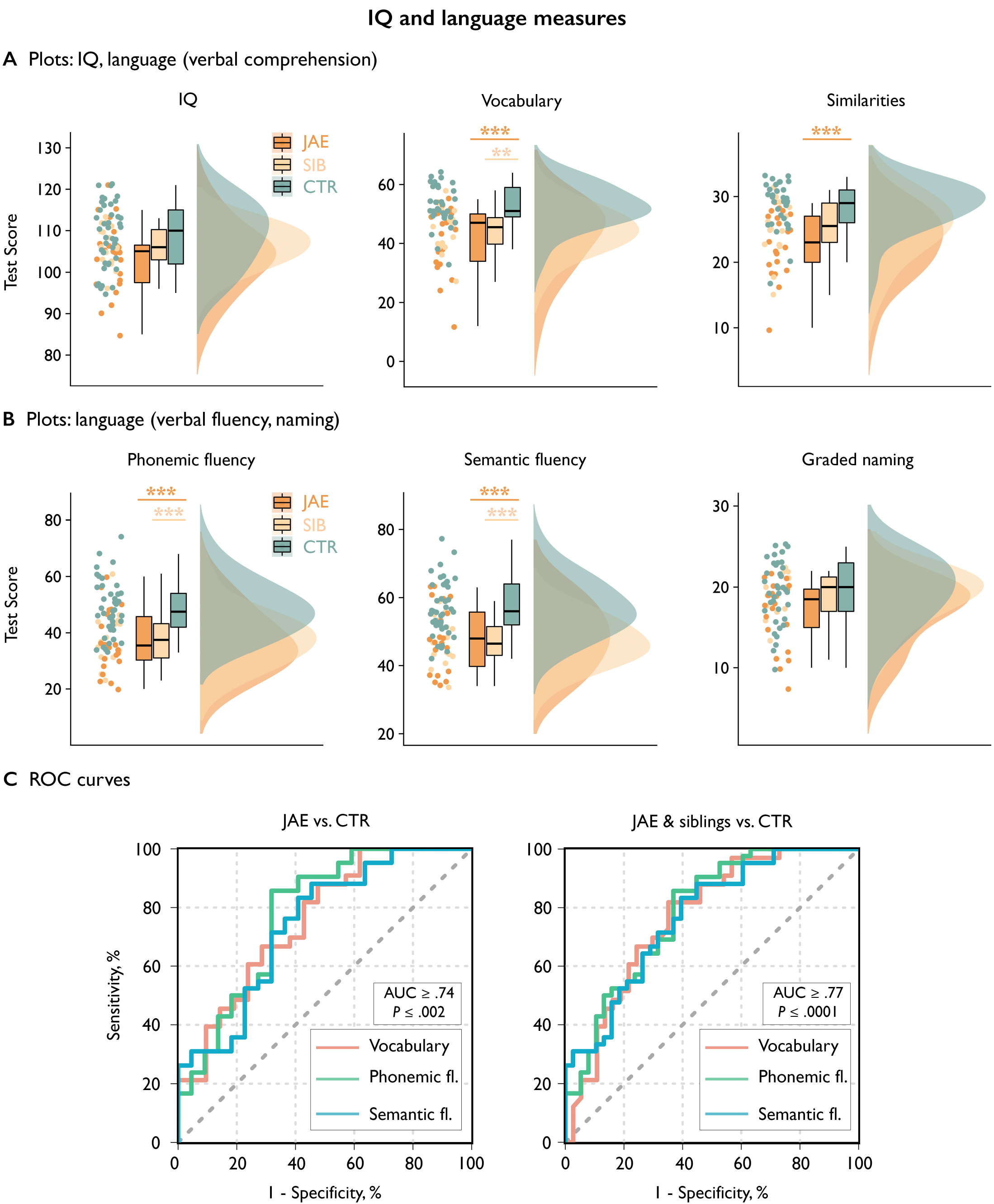
JAE, unaffected JAE Siblings, and controls: IQ and language measures. Panels A and B show neuropsychological test data in individuals with JAE, unaffected JAE siblings (SIB) and controls (CTR) for measures of estimated IQ and verbal comprehension (vocabulary, similarities) (Panel A); and of phonemic fluency, semantic fluency, and naming (Panel B). For each test, we used open-source code to generate *raincloud* plots (https://github.com/RainCloudPlots/RainCloudPlots), and show a combination of raw scores, single datapoints, boxplots, and probability distributions. Statistical details are reported in Table 2 and in the main manuscript text. Asterisks refer to *P-*values for Bonferroni-corrected, age- and sex-adjusted *post-hoc* tests [JAE vs. controls, indicated by underlying orange bars; siblings vs. controls, indicated by underlying sunset (light orange) bars]; **\*\*\*** = *p*<0.001, corrected; **\*\*** = *p*<0.01, corrected; **\*** = *p*<0.05, corrected. Panel C shows receiver-operating characteristic (ROC) curves probing the accuracy with which language measures (vocabulary, phonemic fluency, semantic fluency) can discriminate individuals with JAE from controls (left-sided plots), and a combined group of individuals with JAE and their siblings from controls (right-sided plots). Discrimination was accurate and successful in all instances; all statistical details are provided in the main manuscript text.

**Figure 2.**
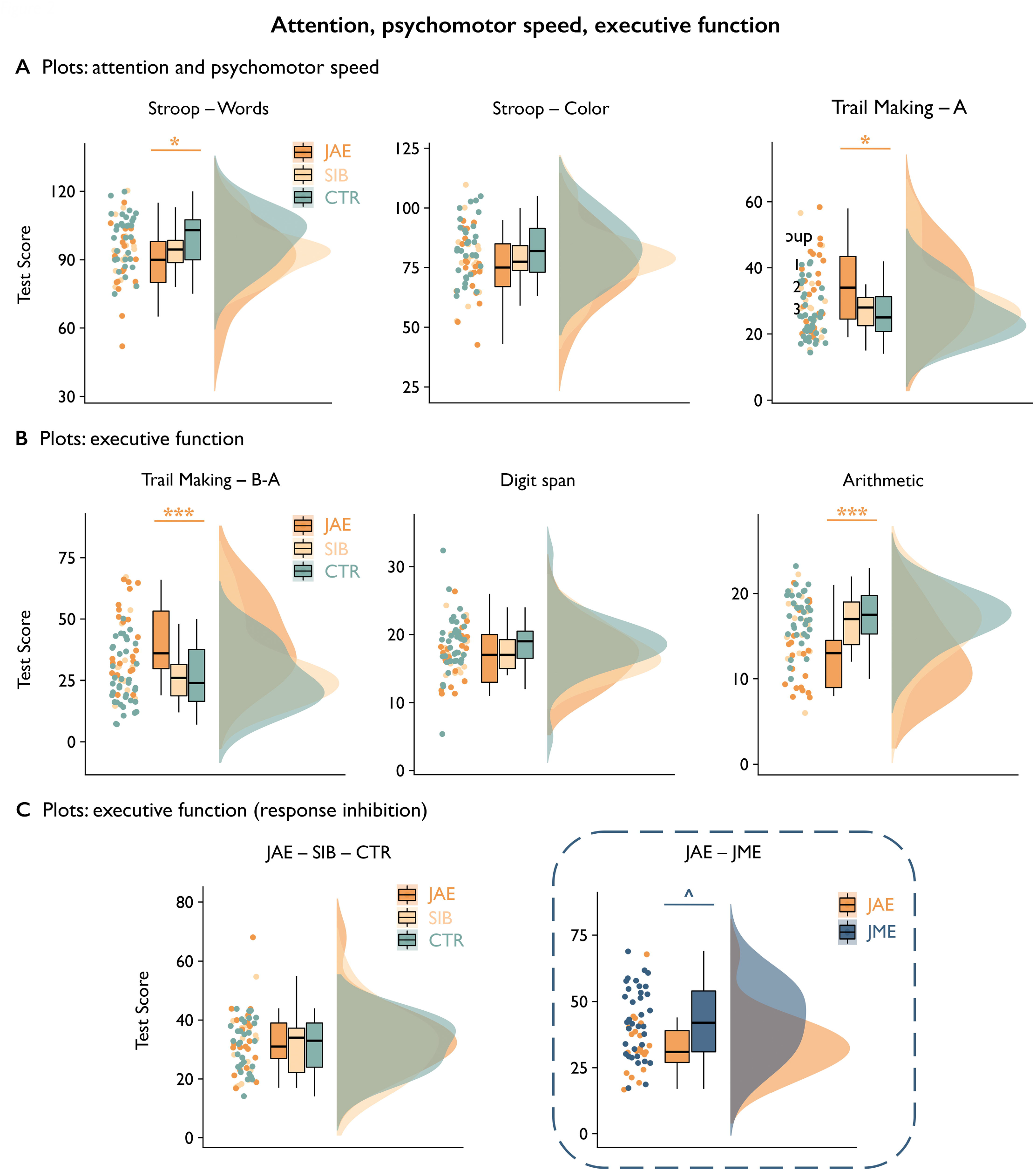
JAE, unaffected JAE siblings, JME, controls: attention, psychomotor speed, executive function. Panels A and B show neuropsychological test data in individuals with JAE, unaffected JAE siblings (SIB) and controls (CTR) for measures of attention and psychomotor speed (Trail Making Test, A, Stroop Word, Stroop Color; Panel A), and executive function (Trail-making B-A, digit span, arithmetic; Panel B). Panel C shows data for a measure of response inhibition (Stroop–– Interference; addressing executive function) in individuals with JAE, unaffected JAE siblings and controls (left-sided plots), and in individuals with JAE juxtaposed to those with JME (right-sided plots, contoured by a rectangular box). For each test, we used open-source code (https://github.com/RainCloudPlots/RainCloudPlots) to generate *raincloud* plots, which show a combination of raw scores, single datapoints, boxplots, and probability distributions. Statistical details are reported in Tables 2 and 3, and in the main manuscript text. Asterisks refer to *P-*values of Bonferroni-corrected, age- and sex-adjusted *post-hoc* tests (JAE vs. controls, indicated by underlying orange bars; JME versus JAE, indicated by underlying blue bar); **\*\*\*** = *p*<0.001, corrected; **\*\*** = *p*<0.01, corrected; **\*** = *p*<0.05, corrected; ^ = *p*<0.05, uncorrected.

**Figure 3.**
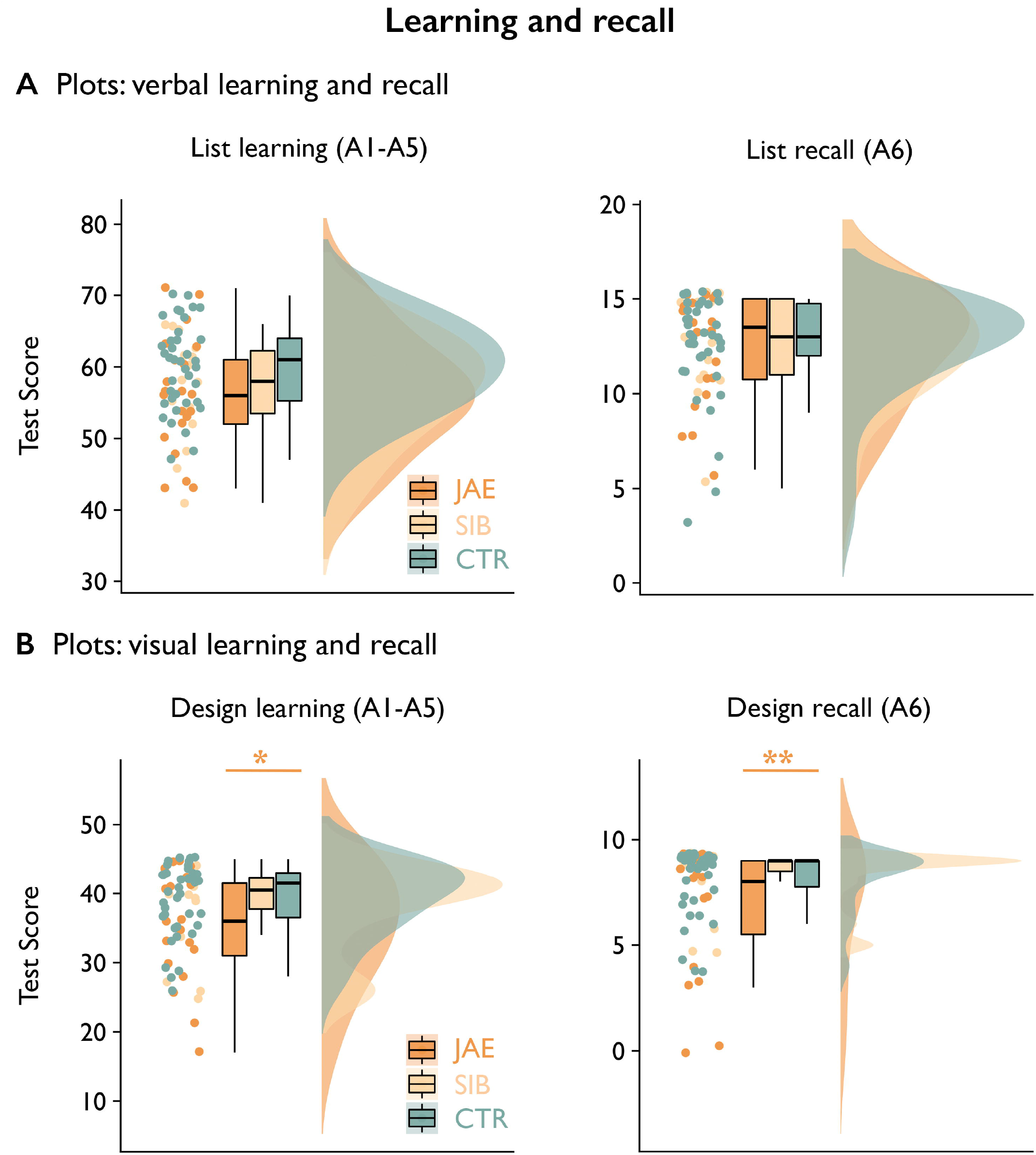
JAE, unaffected JAE Siblings, and controls: verbal and visual learning. Panels A and B show neuropsychological test data in individuals with JAE, unaffected JAE siblings (SIB) and controls (CTR) for measures of verbal learning and recall (Panel A), and of visual learning and recall (Panel B). For each test, we used open-source code to generate *raincloud* plots (https://github.com/RainCloudPlots/RainCloudPlots), which provide a combination of raw scores, single datapoints, boxplots, and probability distributions. Statistical details are reported in Table 2 and in the main manuscript text. Asterisks refer to *P-*values of Bonferroni-corrected, age- and sex-adjusted *post-hoc* tests (JAE vs. controls, indicated by underlying orange bars); **\*\*** = *p*<0.01, corrected; **\*** = *p*<0.05, corrected.

### Individual-level discrimination of JAE and JAE siblings from controls

*Post-hoc* ROC curve analyses (Figure 1C) indicated highly accurate discrimination of patients with JAE from controls via language measures [vocabulary: AUC=.76, standard error (SE)=.07, *p*=.002; phonemic fluency: AUC=.78, SE=.06, *p*=.0003; semantic fluency: AUC=.74, SE=.07, *p*=.001]. Additional analyses of a combined group of patients with JAE and their siblings also showed successful discrimination of patients and siblings from controls, with comparable or higher accuracy (vocabulary: AUC=.77, SE=.06, *p*=.0001; phonemic fluency: AUC=.79, SE=.05, *p*<.0001; semantic fluency: AUC=.78, SE=.05, *p*<.0001). Collectively, these findings show co-segregation of performance on language tests in patients with JAE and their siblings, further corroborating their potential as JAE endophenotypes.

### Sensitivity analyses

Repeat MANCOVA covarying for education in addition to age and sex confirmed a significant effect of group on cognitive performance (*p*=.007); there were no changes in statistical significance and effect size of most tests, particularly those on which JAE patients and siblings showed similar impairment (Supplementary Results). Repeat MANCOVA additionally covarying for self-reported anxiety and depression symptoms yielded a significant effect of group on cognitive performance (*p*=.019); there were no changes in statistical significance and effect size of several tests, particularly those on which JAE patients and siblings demonstrated similar impairment (Supplementary Results).

### Subgroup analysis: JAE patients with ongoing seizures

MANCOVA comparing JAE patients with ongoing seizures against controls showed similar effects as in the main JAE analysis (Wilk’s λ=.28, *F*_(15,16)_=2.77, *p*=.026; Supplementary Table 2). Follow-up analyses showed group differences across multiple cognitive domains, including language (vocabulary, similarities, phonemic and semantic fluency), attention/psychomotor speed (TMT-A, Stroop– Word, Stroop– Color), and executive function (arithmetic, TMT B-A), with similar effect sizes as those of comparisons between the whole JAE group and controls (all *p*_Bonferroni_ ≤.037; *d* range: |0.75| to |1.79|). As distinct from the whole JAE sample, JAE individuals with ongoing seizures had significantly worse naming, visual learning and recall scores than controls (all *p*≤.037, *d=*-.75/-.78/-.80).

### Cognitive performance in JAE versus JME

Statistical details are provided in Table 4. MANCOVA did not show a significant effect of group on cognitive performance (Wilk’s λ44, *F*_(15,24)_=2.08, *p*=.053). Follow-up ANCOVAs showed worse performance on the response inhibition task in JME compared to JAE (*p=*.015, uncorrected; *d=*0.62; Figure 2C), though statistical significance did not survive correction for multiple comparisons across all cognitive tests. For completeness, the cognitive profile of individuals with JME is detailed in Supplementary Table 3. MANCOVA of JME versus controls showed a significant effect of group (Wilk’s λ 35, *F*_(15,30)_=3.77, *p*<.001). Patients with JME performed worse than controls on language (vocabulary, similarities, naming, phonemic fluency), attention/psychomotor speed (TMT-A), executive function (arithmetic, TMT B-A, Stroop– Interference), and (verbal) learning measures (all *p*_Bonferroni_ ≤.019; *d* range: |0.51| to |1.13|).

### Correlation analyses

PCA yielded composite constructs probing the following superordinate domains: *psychomotor speed, language*, and *executive function*. The first PC for *psychomotor speed/language/ executive function* had eigenvalues of 1.84/2.80/2.00 and explained 61.2/56.0/49.9% of the total variance, respectively. Controlling for disease duration, we found significant correlations of age at onset with both *psychomotor speed* and *executive function* (*ρ*=.34, *p*=.014; and *ρ*=.37, *p*=.014, respectively; both *p*_FDR_=.021, adjusted for number of superordinate domains), with later age at onset being associated with better performance for both domains. Correlation of *language* with age at onset was not statistically significant (*ρ*=.12, *p*=.385). Correlations of age and disease duration with *executive function* and *language* did not survive correction for multiple comparisons; complete statistical details are provided in the Supplementary Results.

## DISCUSSION

Despite extensive investigation of JME, CAE or mixed absence epilepsy cohorts,^8,21^ a detailed assessment of the cognitive profile of JAE has been missing. Here we characterized the cognitive phenotype of a homogeneous, well-characterized JAE sample. We identified cognitive impairment spanning multiple domains, including attention and psychomotor speed, language, learning, and executive function. Unaffected JAE siblings were similarly affected as patients on language measures, and showed no differences compared to either patients or controls for the remainder domains. In keeping with previous work in IGE^15^ and in JME specifically,^14,16^ our findings suggest that cognitive impairment in JAE is a familial trait, and may be heritable. Comparison of JME and JAE demonstrated slightly poorer response inhibition in JME, which points to syndrome-specificity of executive function profiles. Similar to JAE, several cognitive domains were also affected in JME, consistent with an overlap of cognitive profiles across IGE syndromes.^1^ Our sensitivity analyses excluded an influence of education and mood symptoms on the reported cognitive signatures. Correlation analyses corroborated the existence of discrete^35^ cognitive phenotypes, i.e., cognitive profiles that are independent of epilepsy syndrome, and are rather influenced by factors such as family history and neurodevelopment. Specifically, an early timing of disease onset may detrimentally affect higher-order cognition, particularly executive function, indicating that neurodevelopmental alterations may be an important determinant of IGE-associated cognitive difficulties.

In our JAE sample, general intellectual abilities were comparable to controls. Previous studies in mixed absence epilepsy reported lower intelligence measures in patients than controls,^8,21^ but we note that such scores fell within a range considered as average for most patients.^8^ We identified poor performance on attention tasks, including sustained, selective and divided attention. Attentional difficulties appear as a key feature of mixed absence epilepsy samples.^8,13,21^ Prior work by Masur *et al*. found attentional deficits in a third of their new-onset CAE cohort, which persisted up to 20 weeks after treatment initiation, irrespective of seizure control.^13^ Collectively, attention deficits may be construed as core characteristic of the cognitive phenotype of JAE, and absence epilepsy more broadly. Interestingly, the above authors also revealed subsequent detrimental effects of attentional deficits on long-term memory, executive function and academic achievement, and other research also pointed to attention as the necessary prerequisite for successful memory and other higher-order abilities.^8,13^ Thus, attentional difficulties may represent an important driver of the multidomain cognitive impairment reported in our and prior studies. Prior combined EEG-functional MRI studies demonstrated altered activity patterns of large-scale brain networks subserving attention, which appeared more prominent during pre-ictal and ictal states.^36,37^ On balance, it is thus possible that attentional difficulties may also be modulated by disease activity.^18,36,37^

Similar to previous studies in mixed absence epilepsy,^21^ executive function was affected in our JAE sample, but not homogeneously, with sparing of response inhibition. Executive function encompasses diverse subprocesses;^38^ here, preserved performance on some executive tests in JAE indicate its non-exclusive contingency on attention, prompting further research on the modulation of specific executive subdomains by attentional difficulties. Secondly, our findings confirm prior evidence of executive dysfunction in mixed IGE samples and in JME,^8^ highlighting commonalities along the IGE spectrum. We also found differences in response inhibition between JME and JAE patients. While the latter finding may be interpreted as exploratory, it indicates that executive function profiles may vary within the IGE spectrum. Our study cannot directly identify the underlying pathophysiology. We speculate that the genetic susceptibility to a specific IGE syndrome, and the associated differences in onset and clinical presentation, may interfere with different circuitry and stages of brain development, which could also influence the cognitive phenotypes. Importantly, response inhibition is elsewhere conceptualized as a marker of impulsivity and poor psychosocial outcome,^39^ which appears more prominent in JME than JAE. Our findings may thus have prognostic implications. We advocate replication with larger samples and more extensive executive function batteries.

Language was affected in the JAE group, in line with previous mixed absence epilepsy studies and meta-analyses.^8,21^ Language abilities appear impaired across the whole epilepsy spectrum, particularly in syndromes with childhood onset. However, the severity of impairment appears slightly greater in absence epilepsies and temporal lobe epilepsy compared to JME and benign epilepsy with centrotemporal spikes.^40^ Language difficulties in syndromes with earlier seizure onset indicates possible neurodevelopmental underpinnings; in CAE, the latter was corroborated by work reporting discordant associations in patients and controls between verbal IQ and structural markers of frontotemporal cortical development.^41^ In our study, we identified similar weaknesses in verbal comprehension and verbal fluency in both people with JAE and their unaffected siblings. As IGE have polygenetic etiology,^42^ investigating unaffected first-degree relatives can identify intermediate phenotypes or *endophenotypes*, i.e. traits that co-segregate in affected families, and help untangle genetic and familial contributions from other variables, such as disease duration or ASM.^17^ The linguistic domain represents a multidimensional construct, shaped by genetic and epigenetic determinants, socio-economic factors, and educational attainment.^43^ Thus, language impairment in JAE may be construed as a *familial* trait, *i*.*e*., a trait that arises from the combination of genetic predisposition, socio-cultural factors, and their interplay. In contrast, the more extensive cognitive difficulties seen in JAE patients than their relatives, as previously documented for IGE and JME, may stem from the additional effects of disease burden, ASM and other factors predisposing to recurrent seizures.^8,14–16^

Frequent seizures, in particular, can undermine cognitive function.^8^ Here, cognitive impairment in the subgroup of JAE patients with ongoing seizures overlapped with that of the whole JAE sample; moreover, effect sizes for language measures with endophenotypic potential were near-identical in the uncontrolled-seizure subgroup, indicating a somewhat limited influence of clinical characteristics. Individuals with JAE and ongoing seizures, however, had more marked difficulties with naming, visual learning, and recall, which rely on mesiotemporal processing. We speculate that neural networks underlying cognitive dysfunction may be broader in those with more severe disease, and more prominently encompass extra-frontal areas, which echoes recent evidence of mesiotemporal alterations in IGE syndromes.^5,44^ We acknowledge, however, that the occurrence of subtle absence seizures in the seizure-free subgroup cannot be completely excluded.

Imaging findings in CAE indicate abnormal frontotemporal cortical geometry^41^ and myelination,^45^ that suggests abnormal neurodevelopment. The timing of disease onset, ranging from late childhood to early adolescence in JAE and JME, may lead to disruption of developmental trajectories in this critical phase, resulting in altered circuit maturation, abnormal cortical topography and, relatedly, cognitive impairment.^46^ In our study, older age at epilepsy onset related to better performance on psychomotor speed and executive function tests. These findings imply that developmental trajectories of slow-maturing frontal networks,^47^ and the higher-order cognitive functions subserved by these, may be affected more markedly in cases with earlier disease onset. It is conceivable that patients with earlier disease onset could accumulate further injury to cognitive networks over time due to chronic disease. In our cohort, however, the effect of age at onset on these cognitive domains was independent of duration, indicating that patterns of cognitive impairment may be established during neurodevelopment. Our finding also align with prior observations by Hermann and colleagues,^35^ who reported cognitive phenotypes in childhood epilepsies that were not syndrome-specific, but influenced by factors linked to brain development, such as age at onset, and spanned different syndromes. Thus, we conclude that early seizures may be more universally harmful to the development of cognitive networks, somewhat irrespective of syndromic classification.

Our study has limitations. Patients did not undergo simultaneous EEG monitoring during neuropsychological testing. While cognitive tests were conducted under the close monitoring of epilepsy specialists, who did not observe clear-cut absence seizures, any potential influence of concurrent subclinical epileptiform discharges on performance could not be formally assessed.^48^ We addressed the potential influence of poorly controlled seizures, but also note that ASM can detrimentally affect cognitive performance, particularly topiramate and zonisamide.^49^ For absence epilepsies specifically, attention deficits appear more frequently associated with sodium valproate use than with other medications.^50^ Here, some individuals with JAE were taking these medications [1 on zonisamide, 1 on topiramate; 10 (43.5%) on sodium valproate], which may have influenced cognition. However, as untreated, unaffected JAE siblings were similarly affected in some domains, such impairment cannot exclusively be attributed to medication effects.

In conclusion, our study characterizes the cognitive profile of JAE, and identifies wide-ranging impairment in language, processing speed, executive function, and memory. Linguistic weaknesses co-segregate in patients with JAE and their unaffected siblings, representing heritable familial traits (endophenotypes). The cognitive profiles of JAE and JME overlap, but there is evidence of syndrome-specific impairment in response inhibition. Cognitive abilities, particularly psychomotor speed and executive function, appear detrimentally modulated by an early seizure onset.

## Abbreviations

AUC: area under the curve
CAE: childhood absence epilepsy
COWAT: Controlled oral word association test;
DEX: Dysexecutive questionnaire;
FDR: false discovery rate;
GTCS: generalized tonic-clonic seizures;
HADS: Hospital anxiety and depression scale;
IGE: idiopathic generalized epilepsy;
IQR: interquartile range;
JAE: Juvenile absence epilepsy;
JME: Juvenile myoclonic epilepsy;
MANCOVA: Multivariate analysis of covariance;
MCAR: missing completely at random;
NART: National adult reading test;
PC(A): Principal component (analysis);
ROC: receiver operating characteristic;
SE: standard error;
WAIS: Wechsler adult intelligence scale.

## ACKNOWLEDGEMENTS

We thank all patients, their siblings, and controls for their participation in this study. Dr. Jonathan O’Muircheartaigh, Professor Veena Kumari, and Professor Mark P. Richardson are acknowledged for their input on previous collaborative projects. Ms. Andrea Hill, Ms. Xandra Harman and Dr. Xin You Tai are acknowledged for their assistance with data acquisition.

## AUTHOR CONTRIBUTIONS

LC, BW and MJK designed the study. MJK and BW supervised the study. LC and BW recruited participants. LC, BW, LAvG, KT, CV, MC, and FX acquired the data. LC, CR, and BW performed the statistical analysis. LC and BW wrote the paper, and revised it based on feedback by CR, FX, KT, CV, MC, LAvG, MJK, JSD, PJT and SB. LC, MJK and BW obtained funding.

## CONFLICT OF INTEREST*/*ETHICAL PUBLICATION STATEMENT

None of the authors have any conflict of interest to disclose.

## CITATION DIVERSITY STATEMENT

Recent work in several fields of science has identified a bias in citation practices such that papers from women and other minority scholars are under-cited relative to the number of such papers in the field (https://github.com/dalejn/cleanBib). Here we sought to proactively consider choosing references that reflect the diversity of the field in thought, form of contribution, gender, race, ethnicity, and other factors. First, we obtained the predicted gender of the first and last author of each reference by using databases that store the probability of a first name being carried by a woman. By this measure (and excluding self-citations to the first and last authors of our current paper), our references contain 12.22% woman(first)/woman(last), 22.47% man/woman, 22.47% woman/man, and 42.83% man/man. This method is limited in that a) names, pronouns, and social media profiles used to construct the databases may not, in every case, be indicative of gender identity and b) it cannot account for intersex, non-binary, or transgender people. Second, we obtained predicted racial/ethnic category of the first and last author of each reference by databases that store the probability of a first and last name being carried by an author of color. By this measure (and excluding self-citations), our references contain 8.17% author of color (first)/author of color(last), 8.81% white author/author of color, 22.15% author of color/white author, and 60.87% white author/white author. This method is limited in that a) names and Florida Voter Data to make the predictions may not be indicative of racial/ethnic identity, and b) it cannot account for Indigenous and mixed-race authors, or those who may face differential biases due to the ambiguous racialization or ethnicization of their names. We look forward to future work that could help us to better understand how to support equitable practices in science.

