## Supplementary Material for "The cognitive phenotype of juvenile absence epilepsy and its heritability: An investigation of patients and unaffected siblings"

Dr Britta Wandschneider, MD PhD

UCL Queen Square Institute of Neurology

Department of Clinical and Experimental Epilepsy

London WC1N 3BG, United Kingdom

### SUPPLEMENTARY METHODS

To ascertain whether letter fluency ought to be assigned to the language or executive function domain, we assessed correlations between letter fluency and (i) other language measures (vocabulary, naming, and semantic fluency), and (ii) other executive function measures [digit span, arithmetic, Trail Making Test (TMT) B-A, Stroop– Interference] across all study participants. For language measures, there were significant correlations between letter fluency and vocabulary ($r$= 0.40, $p$<.001), naming ($r$= 0.25, $p$=.009) and semantic fluency ($r$= 0.49, $p$<.001). On the other hand, letter fluency was significantly correlated with TMT B-A ($r$= -0.28, $p$=.002), digit span ($r$= 0.31, $p$=.001), arithmetic ($r$= 0.35, $p$<.001), but not Stroop– Interference scores ($r$= 0.08, $p$=.44). Consequently, letter fluency was included in the language domain. A similar approach was adopted with regards to the similarities subtest of the WAIS. For language measures, there were significant correlations between similarities and vocabulary ($r$= 0.71, $p$<.001), naming ($r$= 0.34, $p$<.001) and semantic fluency ($r$= 0.35, $p$<.001). Similarities scores were also significantly correlated with TMT B-A ($r$= -0.44, $p$<.001), digit span ($r$= 0.24, $p$=.01), arithmetic ($r$= 0.69, $p$<.001), but not with Stroop interference scores ($r$= -0.13, $p$=.21). Thus, the Similarities WAIS subtest was also included in the language domain.

### SUPPLEMENTARY RESULTS

<u>*MANCOVA on JAE, JAE siblings and controls: sensitivity analyses*</u>

Repeat MANCOVA covarying for *education* in addition to age and sex confirmed a significant effect of group on cognitive performance (Wilk's $\lambda$=.26, $F_{(30,64)}$=2.09, $p$=.007). Significant effects of education were found for estimated IQ, working memory (Digit span), psychomotor speed (Trail Making Test A), and mental flexibility (TMT B-A). Covarying for education affected the statistical significance of group effects for attention and psychomotor speed (TMT A, Stroop–Words), list and design learning (A1-A5), but effect sizes were overall comparable to those of the main analyses. No changes to other test results were observed, particularly those on which individuals with JAE and their unaffected siblings showed similar impairment. Repeat MANCOVA covarying for self-reported *anxiety* and *depression* symptoms in addition to age and sex confirmed a significant effect of group on cognitive performance (Wilk's $\lambda$=.28, $F_{(30,62)}$=1.88, $p$=.019).

Significant effects of anxiety were found for semantic fluency; significant effects of depression were found for design recall (A6). Covarying for education affected significance levels of group effects for attention and psychomotor speed (TMT A, Stroop-Words), design learning (A1-A5) and recall (A6). No changes to other test results were observed, specifically those on which individuals with JAE and their unaffected siblings demonstrated similar impairment.

*Correlation analyses: disease duration and age*

Partial correlations of disease duration with the superordinate cognitive domains of *executive function* and *language* (principal components; see main text), controlling for age at onset, did not survive correction for multiple comparisons ($\rho$=.33/.29, $p$=.031/.037, respectively; both $p_{FDR}$=.056) *executive function* and *language*, but not *psychomotor speed*, were better in older subjects, but these correlations did not survive correction for multiple comparisons ($\rho$=.32, $p$=.037; and $\rho$=.30, $p$=.026, respectively; both $p_{FDR}$=.055; $\rho$=-.003 $p$=.99 for *psychomotor speed*).

### SUPPLEMENTARY TABLE 1. Cognitive tests.

| Domain | Test | Test description |
|---|---|---|
| ***General intellectual abilities*** | *NART* | Participants are asked to read 50 irregularly spelled and/or pronounced British English words. Measure of estimated IQ. |
| ***Attention/Psychomotor Speed*** | *Trail Making Test: Time – Part A* | Participants are asked to connect numbers in ascending order, using a continuous line, as quickly as possible. |
| | *Stroop– Color* | Participants are asked to read a list of named ink colors, and the maximum number of words read within 45 seconds is recorded. |
| | *Stroop– Word* | Participants are asked to read a list of color words, and the maximum number of words read within 45 seconds is recorded. |
| ***Language*** | *Vocabulary (WAIS III)* | Participants are required to provide definitions for 33 specific words of increasing difficulties. |
| | *Similarities (WAIS III)* | Participants are asked to describe the relationship (common link) between 19 pairs of words. |
| | *Controlled Oral Word Association Test* | Participants are asked to name words starting with a specific letter (F, A, or S) in one minute. Probes letter fluency. |
| | *Category fluency test* | Participants are asked to name items subsumed under a specific category in one minute; in our study, the three tested categories were: animals, fruits, and vegetables. |
| | *McKenna Graded Naming test* | Participants are asked to name 30 items of graded difficulty that are depicted as black-and-white line drawings. |
| ***Learning*** | *AMIPB: List Learning* | Participants are asked to memorize a 15-item word list over five trials (List A1-A5) |
| | *AMIPB: List Recall* | Participants are asked to recall the original word list, memorized during the learning phase of the test, following a distracting list. |
| | *AMIPB: Design Learning* | Participants are asked to reproduce a 9-element design on a 4 × 4 grid over five consecutive trials (Design A1-A5). |
| | *AMIPB: Design Recall* | Participants are asked to reproduce the original design, memorized during the learning phase of the test, following a distracting design |
| ***Executive function*** | *Digit Span* | Participants are asked to repeat a set of numbers of increasing length in the correct order upon presentation; then, they are asked to repeat second set of numbers in reverse order. Probes working memory. |
| | *Arithmetic* | Participants are asked to solve orally presented arithmetic problems without using pen and paper. Probes working memory. |
| | *Trail Making Test: Task Switching (B-A)* | Participants are asked to connect numbers and letters of the alphabet in sequence, alternating between letters and numbers, as fast as possible. Probes cognitive flexibility. |
| | *Stroop – Interference* | Participants are asked to name the ink color of color words that written with an incongruent color as quickly as possible. Probes response inhibition. |

*Abbreviations.* AMIPB= Adult Memory and Information Processing Battery; NART= National Adult Reading Test; WAIS= Wechsler Adult Intelligence Scale. Test references are provided in the main manuscript text.

### SUPPLEMENTARY TABLE 2. Comparison of JAE with ongoing seizures and controls.

| **Multivariate model.** Wilk's lambda=.28, $F_{(15,16)}$=2.77, ***p*= .026** | | | | |
|---|---|---|---|---|
| | **Effect of group (*F* statistic)** | **$P_{FDR}$ value (uncorr. *P*)** | **Mean (SD)** | **Effect size (Cohen's *d*)** |
| Estimated IQ (NART) | $F_{1,53}$ = 4.31 | .0562 (.043) | JAE: 103.1 (9.0)<br>CTR: 108.8 (7.7) | JAE vs CTR: **-.60** |
| Vocabulary | $F_{1,43}$ = 10.46 | **.0043 (.002)** | JAE: 43.5 (10.1)<br>CTR: 52.6 (7.0) | JAE vs CTR: **-1.04** |
| Similarities | $F_{1,43}$= 15.81 | **.001 (.0003)** | JAE: 23.5 (4.2)<br>CTR: 28.7 (3.6) | JAE vs CTR: **-1.27** |
| McKenna Graded Naming | $F_{1,49}$ = 5.40 | **.0368** (.024) | JAE: 16.1 (3.6)<br>CTR: 19.5 (4.0) | JAE vs CTR: **-.70** |
| Phonemic Fluency (F, A, S; sum of all words per letter) | $F_{1,53}$ = 9.66 | **.0057 (.003)** | JAE: 38.2 (12.5)<br>CTR: 48.6 (9.8) | JAE vs CTR: **-.93** |
| Semantic Fluency (Animals, fruits, vegetables, sum of all items per category) | $F_{1,53}$ = 16.03 | **.001 (.0002)** | JAE: 47.2 (11.1)<br>CTR: 57.1 (8.1) | JAE vs CTR: **-1.18** |
| Digit Span | $F_{1,49}$ = 2.04 | .180 (.159) | JAE: 17.0 (4.3)<br>CTR: 18.7 (4.3) | JAE vs CTR: -.45 |
| Arithmetic | $F_{1,43}$ = 29.96 | **<.0001 (.000002)** | JAE: 11.7 (4.0)<br>CTR: 17.1 (3.0) | JAE vs CTR: **-1.79** |
| Trail Making Test (A) (seconds) | $F_{1,51}$ = 11.45 | **.0024 (.001)** | JAE: 35.2 (11.3)<br>CTR: 26.2 (7.7) | JAE vs CTR: **1.02** |
| Trail Making Test (B-A) (seconds) | $F_{1,50}$ = 12.25 | **.0024 (.001)** | JAE: 40.2 (14.1)<br>CTR: 26.2 (12.0) | JAE vs CTR: **1.06** |
| Stroop - Words (items in 45 seconds) | $F_{1,41}$ = 7.20 | **.017 (.010)** | JAE: 89.8 (14.8)<br>CTR: 99.3 (11.5) | JAE vs CTR: **-.87** |
| Stroop - Colour (items in 45 seconds) | $F_{1,41}$ = 5.37 | **.0368** (.026) | JAE: 75.5 (13.4)<br>CTR: 83.0 (12.1) | JAE vs CTR: **-.75** |
| Stroop Interference (Words + Colour)/2 minus items in Colored Words) | $F_{1,41}$ = .55 | .465 (.465) | JAE: 33.9 (12.7)<br>SIB: 31.3 (10.5)<br>CTR: 31.6 (8.5) | JAE vs CTR: .24 |
| List Learning (A1-A5) | $F_{1,48}$ = 3.43 | .085 (.070) | JAE: 56.3 (9.2)<br>CTR: 60.1 (6.1) | JAE vs CTR: -.58 |
| List Recall (A6) | $F_{1,47}$ = .94 | .3581 (.337) | JAE: 12.0 (23.3)<br>CTR: 12.5 (2.8) | JAE vs CTR: -.32 |
| Design Learning (A1-A5) | $F_{1,49}$ = 15.86 | **.001 (.0002)** | JAE: 32.7 (8.4)<br>CTR: 39.3 (5.2) | JAE vs CTR: **-1.27** |
| Design Recall (A6) | $F_{1,47}$ = 14.87 | **.001 (.0003)** | JAE: 6.0 (3.5)<br>CTR: 8.1 (1.5) | JAE vs CTR: **-1.23** |

*Abbreviations.* CTR = Controls; JAE = Juvenile Absence Epilepsy; NART= National Adult Reading Test; SD= standard deviation; uncorr.= uncorrected. All statistical analyses controlled for age and sex.

### SUPPLEMENTARY TABLE 3. Comparison of JME and controls.

| ***Multivariate model***: Wilk's Lambda= .35, $F_{(15,30)}$=3.77, ***p< .001*** | | | | |
|---|---|---|---|---|
| | **Effect of group (*F* statistic)** | ***P<sub></sub>*** $P_{FDR}$ **value (uncorr. *P*)** | **Mean (SD)** | **Effect size (Cohen's *d*)** |
| Premorbid IQ | $F_{1,72}$ = .37 | .661 (.544) | JME: 108.4 (10.9)<br>CTR: 108.8 (7.7) | JME vs CTR: -.13 |
| Vocabulary | $F_{1,66}$ = 11.71 | **.003 (.001)** | JME: 48.1 (9.5)<br>CTR: 52.6 (7.0) | JME vs CTR: **-.75** |
| Similarities | $F_{1,66}$ = 27.56 | **<.0001 (<.0001)** | JME: 23.6 (4.4)<br>CTR: 28.7 (3.6) | JME vs CTR: **-1.13** |
| McKenna | $F_{1,72}$ = 7.37 | **.019 (.008)** | JME: 18.3 (3.9)<br>CTR: 19.5 (4.0) | JME vs CTR: **-.59** |
| Phonemic Fluency<br>(F, A, S; sum of all words per letter) | $F_{1,77}$ = 14.16 | **.002 (.0003)** | JME: 41.1 (11.08)<br>CTR: 48.6 (9.8) | JME vs CTR: **-.79** |
| Semantic Fluency<br>(Animals, fruits, vegetables, sum of all items per category) | $F_{1,77}$ = 2.39 | .179 (.126) | JME: 52.2 (12.4)<br>CTR: 57.1 (8.1) | JME vs CTR: -.33 |
| Digit Span | $F_{1,72}$ = .13 | .822 (.725) | JME: 19.0 (4.4)<br>CTR: 18.7 (4.3) | JME vs CTR: -.08 |
| Arithmetic | $F_{1,63}$ = 13.66 | **.002 (.0005)** | JME: 14.3 (4.2)<br>CTR: 17.1 (3.0) | JME vs CTR: **-.79** |
| Trail Making Test (A)<br>(seconds) | $F_{1,75}$ = 5.80 | **.034 (.018)** | JME: 31.3 (10.3)<br>CTR: 26.2 (7.7) | JME vs CTR: **.51** |
| Trail Making Test (B-A)<br>(seconds) | $F_{1,74}$ = 11.48 | **.003 (.001)** | JME: 37.4 (16.9)<br>CTR: 26.2 (12.0) | JME vs CTR: **.71** |
| Stroop Words<br>(items in 45 seconds) | $F_{1,62}$ = .02 | .899 (.899) | JME: 99.7 (20.2)<br>CTR: 99.3 (11.5) | JME vs CTR: -.03 |
| Stroop Colour<br>(items in 45 seconds) | $F_{1,62}$ = 2.58 | .175 (.113) | JME: 74.3 (14.9)<br>CTR: 83.0 (12.1) | JME vs CTR: -.37 |
| Stroop Interference<br>(Words + Colour)/2 minus items in Colored Words) | $F_{1,62}$ = 11.69 | **.0009 (.0001)** | JME: 43.0 (13.4)<br>CTR: 31.6 (8.5) | JME vs CTR: **-.79** |
| List Learning (A1-A5) | $F_{1,73}$ = 7.29 | **.019 (.009)** | JME: 54.2 (8.6)<br>CTR: 60.1 (6.1) | JME vs CTR: **-.57** |
| List Recall (A6) | $F_{1,73}$ = .07 | .846 (.796) | JME: 11.6 (2.6)<br>CTR: 12.5 (2.8) | JME vs CTR: -.06 |
| Design Learning (A1-A5) | $F_{1,75}$ = 4.77 | .054 **(.032)** | JME: 34.8 (8.9)<br>CTR: 39.3 (5.2) | JME vs CTR: -.47 |
| Design Recall (A6) | $F_{1,75}$ = .96 | .432 (.329) | JME: 7.4 (2.0)<br>CTR: 8.1 (1.5) | JME vs CTR: -.21 |

*Abbreviations.* CTR= healthy controls; JME= Juvenile Myoclonic Epilepsy; NART= National Adult Reading Test; SD= standard deviation; uncorr.= uncorrected. All statistical analyses controlled for age and sex.
